# Effect of an Integrated Maternal and Neonatal Health Intervention on Maternal Healthcare Utilization addressing Inequity in Rural Bangladesh

**DOI:** 10.1101/2022.02.27.22271594

**Authors:** Anisuddin Ahmed, Nafisa Lira Huq, Fariya Rahman, Tania Sultana Tanwi, Md. Abu Bakkar Siddique, Aniqa Tasnim Hossain, Saraban Tahura Ether, Ema Akter, Tazeen Tahsina, Shams El Arifeen, Ahmed Ehsanur Rahman

**Affiliations:** International Centre for Diarrheal Disease Research, Bangladesh (icddr,b). 68, Shaheed Tajuddin Ahmed Sarani, Mohakhali, Dhaka 1212, Bangladesh

**Keywords:** Maternal Health, Child Health, Timely ANC Visit, ANC Checkup, Skilled Delivery, Timely PNC, Skilled Health Care Provider, Bangladesh

## Abstract

**Background:** Although Bangladesh has made significant improvements in maternal, neonatal, and child health, the disparity between rich and poor remains a matter for concern.

**Objective:** The study aimed to increase coverage of women in seeking skilled maternal healthcare services while minimizing inequity gap among different socioeconomic groups.

**Methods:** icddr, b implemented an integrated maternal and neonatal health (MNH) intervention between 2009 and 2012, in Shahjadpur sub-district of Shirajganj district, Bangladesh. The study was pre- and post-test in design for evaluation including baseline and endline surveys. The baseline and endline surveys were conducted among 3158 and 3540 recently delivered mothers respectively. Asset index derived from household assets using principal component analysis was categorized into five ordinal categories, i.e. Poor, Less poor, Middle, Upper middle, Rich. Inequity in maternal healthcare utilization was calculated for the baseline and endline periods using rich-to-poor ratio and the concentration index.

**Result:** Mean age of mothers were 23.5 and 24.3 years in baseline and endline, respectively. Reduction in rich-poor ratio was quite large in utilization of skilled 4+ antenatal care (ANC) (2.4:1 to 1.1:1), childbirth (1.7:1 to 1.0:1), and postnatal care (PNC) (2.5:1 to 1.0:1) from trained providers between these two surveys. The concentration indices (CI) in endline for skilled 4+ ANC (CI: 0.220 and 0.013), delivery (CI: 0.161 and -0.021), and PNC (CI: 0.197 and -0.004) were found to be lower than the indices in baseline period respectively.

**Conclusion:** The MNH intervention was successful in reducing inequity in receiving skilled 4+ ANC, delivery, and PNC in rural Bangladesh. Improvements in maternal healthcare utilizations by poor mothers would be influenced by the properly designed and integrated demand- and supply-side MNH interventions package.

## Introduction

Universal Health Coverage (UHC)-a global agenda aims to cover health services at a reasonable cost as per individual’s requirements (1). Hence, Maternal, Neonatal, and Child Health (MNCH) services e.g., antenatal care (ANC), skilled delivery, and postnatal care (PNC) are three MNCH services covered under UHC package (2). ANC, skilled delivery, and PNC are considered effective means to prevent pregnancy, delivery, and postnatal complications and thus mean to reduce mortality (3, 4). Despite having UHC scheme, many countries around the world are still struggling to narrow down the service gap among the service recipients of rich and poor quintile (5). In 2015, the State of Inequity report comprised data on MNCH inequity-prone 86 low- and middle-income countries (5). The report showed that globally, the richest women can avail skilled delivery more than the poorest women by more than 80% points and, almost half of the reported countries showed a difference of 25% points among the richest and the poorest women in ANC uptake (5). The global activities to reduce maternal mortality and other adverse maternal health outcomes is not inclusive of achieving equity. It therefore is important to ensure access to essential maternal, neonatal, and child healthcare services by women and children, irrespective of their household socioeconomic status to achieve targets towards the SDGs (6).

With persisting socioeconomic differences, the inequity scenario in Bangladesh is quite similar to this global finding. Bangladesh Demographic and Health Survey 2017-18 measured that the skilled ANC seeking percentage between the richest and the poorest quintile differ by 34% percentage points (97.2% vs 63.6%) (7). Likewise, health facility delivery is higher among the women of the highest wealth quintile in comparison to the lowest wealth quintile (77.9% vs 26.3%) and similarly, rich and poor gap is also observed in terms of PNC as well (7). According to BDHS 2017, almost 71.5% mothers from the lowest wealth quintile do not avail PNC service at all and for the highest wealth quintile this percentage is 17.9% (7).

To act upon service gaps and improve MNCH status, since 2000 several governmental and non-governmental programs are functional in Bangladesh. The programs include Maternal Health Voucher Scheme, Emergency Obstetrical Care Services (EmOC) and Government’s nationwide community skilled birth attendants (CSBAs) training program and midwifery training program to increase coverage of skilled provider’s care during pregnancy to ensure a healthy and secured birth outcome for women in general (8, 9). However, insufficient implementations of these efforts might contribute to the continuing high maternal mortality of the country (10, 11).

Hence, an integrated evidence-based MNH intervention package was implemented by the International Centre of Diarrheal Disease Research, Bangladesh (icddr, b) in partnership with Bangladesh government over 4 years in a rural sub-district, to strengthen the existing healthcare system. The package contained birth and newborn care preparedness counselling, updated safe delivery-kit, management of postpartum hemorrhage (PPH) through routine implementation of active management of third stage labor (AMTSL), misoprostol, and safe blood transfusion, management of eclampsia by MgSO_4_ and home-based essential newborn care by community skilled birth attendants (CSBAs) for sensitizing the mother, formation of community support groups (CSGs) for community sensitization (12). This package has been tested and evaluation revealed the underprivileged women benefitted from skilled pregnancy care (12). This paper aims to understand the role of the skilled healthcare providers on inequity in the utilization of skilled health care services during pregnancy, childbirth, and postnatal period.

## Materials and methods

### Study design and settings

The study was quasi-experimental in design. The baseline and endline surveys were conducted to evaluate the effect of the intervention. The integrated intervention package was introduced in Shahjadpur, a sub-district of Shirajganj district under the Rajshahi division, Bangladesh covering about 600,000 populations, between 2009 and 2012.

### Sampling and study participants

One municipality area and thirteen unions of Shahjadpur were divided into clusters. Each cluster accommodated approximately 3,000 people. Two hundred clusters were made out of a total population of 583,350. In baseline 80 clusters (interviewed 3158 mothers) and in endline 100 clusters (interviewed 3540 mothers) were randomly selected. Data collection methodology and survey questionnaire were the same in both the surveys. Sample size estimation was based on the reduced neonatal mortality (from 37 to 21 per 1,000 live births after 2-years of completion of the intervention) with 95.0% confidence and 80.0% power, a minimum of 1,250 mothers who experienced live births were estimated for each of the surveys. A total of 1,290 mothers were needed to conduct analysis for the stillbirth outcome also (assuming 30/1,000 births). Compensating the non-response at 5%, a total of 1,360 mothers who had an experience of childbirth after 28 weeks of gestation were required. Finally, a total of at least 2,720 recently delivered mothers (delivery occurred in the last 6 months) were estimated in each baseline and endline survey using the design-effect of 2 for cluster sampling. Married women of reproductive age of 15-49 years who had a delivery outcome in the last 6-months before the date of interviews were eligible.

### Data collection

The baseline and endline surveys were conducted in 2009 and 2012 respectively. The survey questionnaire included possession of household assets, demographic characteristics including age, education, care-seeking patterns for antenatal/natal/postnatal periods, last birth outcomes, newborn care, and neonatal care. Questions of the questionnaire were guided by the compendium of Maternal and Newborn Health Tools developed by MEASURE Evaluation (13). The questionnaire was finalized after adjusting the contextual thematic aspects for Bangladesh that emerged from the pre-testing of the questionnaire on completion of the training of the data collectors. The data collection team comprised of four field research supervisors (FRSs) and 16 Field Research Assistants (FRAs). The FRAs were responsible for the structured interview at the household level, the quality of the interview was ensured by close monitoring by the FRSs. All the eligible women at each block were identified by a door-to-door visit and approached for the interview.

### Data management and quality assurance

An efficient research team led by an experienced leader was involved in the data collection. To maintain the quality of data, the team leader supervised the data collection team and was responsible for spot verification of the completeness of every interview. Furthermore, a research investigator (RI) and a project research physician (PRP) were appointed to coordinate the data collection team on a daily and weekly basis for checking the data quality. To ensure the accuracy and data validity, PRP and RI conducted re-interview in a significant amount. Simultaneously, an expert programmer team of the Maternal and Child Health Division (MCHD) of icddr, b designed a database template using Dot net (Version-10) software to enter all the data online. The data template housed an advanced design to avoid any scope of variables missing. Skipping options were also maintained strictly and logically to avoid entry mistakes. The expert data management team entered all the pre and post-coded data through an online database simultaneously. For post coding of data, the data management team cooperated with the research team.

### Ethics

This study did possess no more than minimal risk on the study participants. All the study participants were persuaded to obtain written informed consent before the interviews were conducted. The research team sought approval from the Institutional Review Board (IRB) of icddr, b before data collection in the field. Since the study only enrolled married women, consent was acquired from the husband in the case of a woman under the age of 18.

### Data analysis

Data analysis was done using STATA 13.1 (Stata, College Station, TX, USA). Chi-square test of independence was used to test the crude association between the different maternal health indicators and the surveys (baseline and endline) by the socioeconomic status of the respondents. Further, principal component analysis was done to measure respondents’ socioeconomic status by a wealth index derived from the household assets (14). The assets included housing materials (e.g. type of roof), housing facilities (e.g. sources of drinking water, types of toilet), durable consumption goods (e.g. television, bicycle, watch, table, chair), and land ownership. The wealth index was then categorized into five ordinal categories including poor, less poor, middle, upper middle, and rich. The concentration indices for inequity measurement were calculated for utilization of each maternal health care-seeking indicators and wealth scores against each respondent. The indicators and scores were plotted to generate the concentration curves to observe the changes in inequity between the baseline and the endline time period.

### The operational definition of concentration curves and index

The concentration curve delineates inequity by plotting the cumulative percentage of health variable with respect to the cumulative percentage of the population ranked from the poor to the rich. When the concentration curve is conformed to the line of equity at 45°, it then shows the perfect equity. The curve lies above the perfect equity line means the health variable is more concentrated among the poor and vice versa. The concentration index gives the magnitude of inequity which is ranged from -1 to +1 and is also defined as twice the area between the concentration curve and the line of equity. Perfect equity is achieved when the index value is zero; the index value closer to -1 means the disproportionate concentration of health variable increases among the poor whereas the disproportionate concentration of health variable increases among the rich if the index value gets closer to +1 (15).

Maternal healthcare-seeking indicators included in this paper were ANC and PNC by a skilled provider and delivery by a skilled birth attendant. These indicators are binary variables and that faces a problem with the standard concentration index which not necessarily always be within the range of -1 to +1 (16). Wagstaff developed a concentration index modified by re-scaling the standard index to keep unscathed the relative inequity variance property of the concentration index (17). For the corrected concentration index for this study, *conindex* command of STATA has been used (18).

## Results

**“Table 1”** describes most of the demographic characteristics of the mothers during the baseline and endline survey periods were not significant in differences across the five categories of the wealth index. Overall, one-fifth of the mothers in both the baseline and the endline surveys were aged below 20 years across the five wealth index categories. Though there were significant differences in education level among the mothers who belonged in the wealth index groups during the baseline period, no significant differences were found among the groups at the endline period. Overall, more than 10.0% of the mothers in both the survey periods had completed eight or more years of schooling. Significant differences were found at their husbands’ education level among the different wealth index groups during both the baseline and the endline surveys. Overall, 40.0% of their husbands had no formal education during the surveys. Almost all the mothers were homemakers and two-thirds of the mothers experienced multi-para during the surveys.

**Table 1:** Sociodemographic characteristics of recently delivered mothers.

| Sociodemographic characteristics | Baseline (% of mothers) |  |  |  |  | Endline (% of mothers) |  |  |  |  |
| --- | --- | --- | --- | --- | --- | --- | --- | --- | --- | --- |
|  | Poor | Less poor | Middle | Upper middle | Rich | Poor | Less poor | Middle | Upper middle | Rich |
|  | n= 631 | n= 635 | n= 629 | n= 632 | n= 631 | n= 714 | n=707 | n= 703 | n= 706 | n= 710 |
| Maternal age (in years) |  |  |  |  |  |  |  |  |  |  |
| ≤19 | 20.8 | 22.7 | 22.6 | 23.4 | 23.8 | 19.9 | 22.4 | 19.6 | 22.5 | 20.1 |
| 20-29 | 65.0 | 61.7 | 60.1 | 62.8 | 63.2 | 60.9 | 58.7 | 62.0 | 59.8 | 61.3 |
| ≥30 | 14.3 | 15.6 | 17.3 | 13.8 | 13.0 | 19.2 | 19.0 | 18.4 | 17.7 | 18.6 |
| *p-value* | 0.454 |  |  |  |  | 0.858 |  |  |  |  |
| Maternal education level (in years) |  |  |  |  |  |  |  |  |  |  |
| No schooling | 46.1 | 40.0 | 35.5 | 30.5 | 28.1 | 33.6 | 28.9 | 32.2 | 30.7 | 31.6 |
| 1-5 | 30.1 | 32.4 | 34.3 | 34.7 | 34.2 | 37.5 | 36.8 | 32.7 | 36.2 | 35.8 |
| 6-7 | 14.6 | 15.9 | 18.9 | 21.2 | 20.1 | 18.9 | 21.6 | 20.2 | 19.5 | 18.1 |
| ≥8 | 9.2 | 11.7 | 11.3 | 13.6 | 17.6 | 9.9 | 12.7 | 14.9 | 13.6 | 14.5 |
| *p-value* | 0.000 |  |  |  |  | 0.161 |  |  |  |  |
| Husband education level (in years) |  |  |  |  |  |  |  |  |  |  |
| No schooling | 52.5 | 47.6 | 41.5 | 38.0 | 34.2 | 42.0 | 43.0 | 42.5 | 42.2 | 40.9 |
| 1-5 | 24.1 | 22.1 | 27.8 | 25.5 | 27.3 | 28.9 | 24.9 | 25.3 | 26.1 | 27.4 |
| 6-7 | 8.4 | 11.0 | 11.8 | 13.3 | 11.4 | 12.5 | 13.6 | 12.7 | 10.4 | 8.9 |
| ≥8 | 15.1 | 19.4 | 18.9 | 23.3 | 27.1 | 16.7 | 18.5 | 19.5 | 21.3 | 22.9 |
| *p-value* | 0.000 |  |  |  |  | 0.059 |  |  |  |  |
| Occupation |  |  |  |  |  |  |  |  |  |  |
| Homemaker | 94.0 | 95.1 | 94.6 | 95.4 | 94.5 | 93.0 | 91.0 | 91.5 | 91.2 | 92.5 |
| Others | 6.0 | 4.9 | 5.4 | 4.6 | 5.6 | 6.7 | 8.8 | 8.4 | 8.4 | 7.5 |
| *p-value* | 0.808 |  |  |  |  | 0.648 |  |  |  |  |
| Reproductive History |  |  |  |  |  |  |  |  |  |  |
| Primi-para | 32.3 | 32.0 | 31.3 | 39.4 | 37.6 | 27.5 | 33.1 | 30.3 | 32.0 | 33.1 |
| Multi-para | 67.7 | 68.0 | 68.7 | 60.6 | 62.4 | 72.6 | 66.9 | 69.7 | 68.0 | 66.9 |
| p-value | 0.000 |  |  |  |  | 0.110 |  |  |  |  |

**“Table 2”** draws a comparison between the baseline and endline estimates of antenatal, delivery, and postnatal care services with skilled providers showed significant increases among the mothers of the five wealth quintiles. At the baseline survey period, 5.9% and 14.0% of the mothers in the poor and rich groups received more than four ANC services from the skilled providers respectively and at the endline period, more than one-third of the mothers received more than four ANC services across each of the wealth index groups. Between the two survey periods, it was found that the differences in receiving 4+ANC services significantly increased across the wealth index. Substantial differences were found for using of skilled birth attendants between the baseline and the endline periods across the poor to the rich groups. About one-fifth (20.6%) of the mothers in the poor group delivered by the skilled birth attendant during the baseline period which has been significantly increased to 45.0% during the endline surveys. A similar change has been observed (35.3% vs. 43.7%) among the mothers in the rich group between the surveys. Use of skilled PNC services was extremely low before the intervention in both the poor and the rich groups (2.4% and.5.9% respectively) and after the intervention, there was a significant improvement in both the quintiles (24.9% and 25.9% respectively). The inequity in the utilization of more than four skilled ANC services, skilled childbirth, and skilled PNC services declined significantly over the two time periods of the baseline and endline. The reduction in the rich-poor ratio was quite large in the utilization of skilled 4+ANC services which was reduced from 2.4:1 to 1.1:1 between the baseline and the endline periods.

**Table 2:** Maternal health care-seeking behavior by their socioeconomic status during the baseline and the endline.

| MNH care services | % of mothers |  |  |  |  |  |
| --- | --- | --- | --- | --- | --- | --- |
|  | Poor/ | Less poor/ | Middle/ | Upper middle/ | Rich/ | Rich-Poor ratio |
|  | Poorest | Poorer | Poor | Less poor | Least poor |  |
|  | % (n) | % (n) | % (n) | % (n) | % (n) |  |
| 4+ANC by skilled providers |  |  |  |  |  |  |
| Baseline | 5.9 | 6.6 | 7.6 | 10.8 | 14.0 | 2.4:1 |
| End line | 31.5 | 34.9 | 30.6 | 30.9 | 35.5 | 1.1:1 |
| *p-value* | 0.000 | 0.000 | 0.000 | 0.000 | 0.000 |  |
| Delivery by skilled providers |  |  |  |  |  |  |
| Baseline | 20.6 | 21.1 | 26.4 | 29.0 | 35.3 | 1.7:1 |
| End line | 45.0 | 47.1 | 45.7 | 45.2 | 43.7 | 1.0:1 |
| *p-value* | 0.000 | 0.000 | 0.000 | 0.000 | 0.002 |  |
| PNC by skilled providers |  |  |  |  |  |  |
| Baseline | 2.4 | 3.0 | 3.3 | 3.0 | 5.9 | 2.5:1 |
| End line | 24.9 | 24.9 | 27.5 | 23.7 | 25.9 | 1:1 |
| *p-value* | 0.000 | 0.000 | 0.000 | 0.000 | 0.000 |  |

**“Figure 1”** shows across the five wealth index groups, the proportion of mothers received more skilled 4+ANC services during the endline period as compared to the baseline period. However, the mothers among the poor (4.9% vs. 22.1%), less poor (5.8% vs. 22.9%), and middle (5.6% vs. 20.9%) groups received 4 times higher-skilled 4+ANC services at the facilities during the endline period as compared to the baseline period respectively. This proportion also significantly increased for CSBAs as a source of ANC services during endline as compared to the baseline surveys. A higher proportion reported for the facility ANC services (more than one-fifth) in comparison to ANC from CSBAs at home (more than 6%) at the endline period.

**Figure 1:**
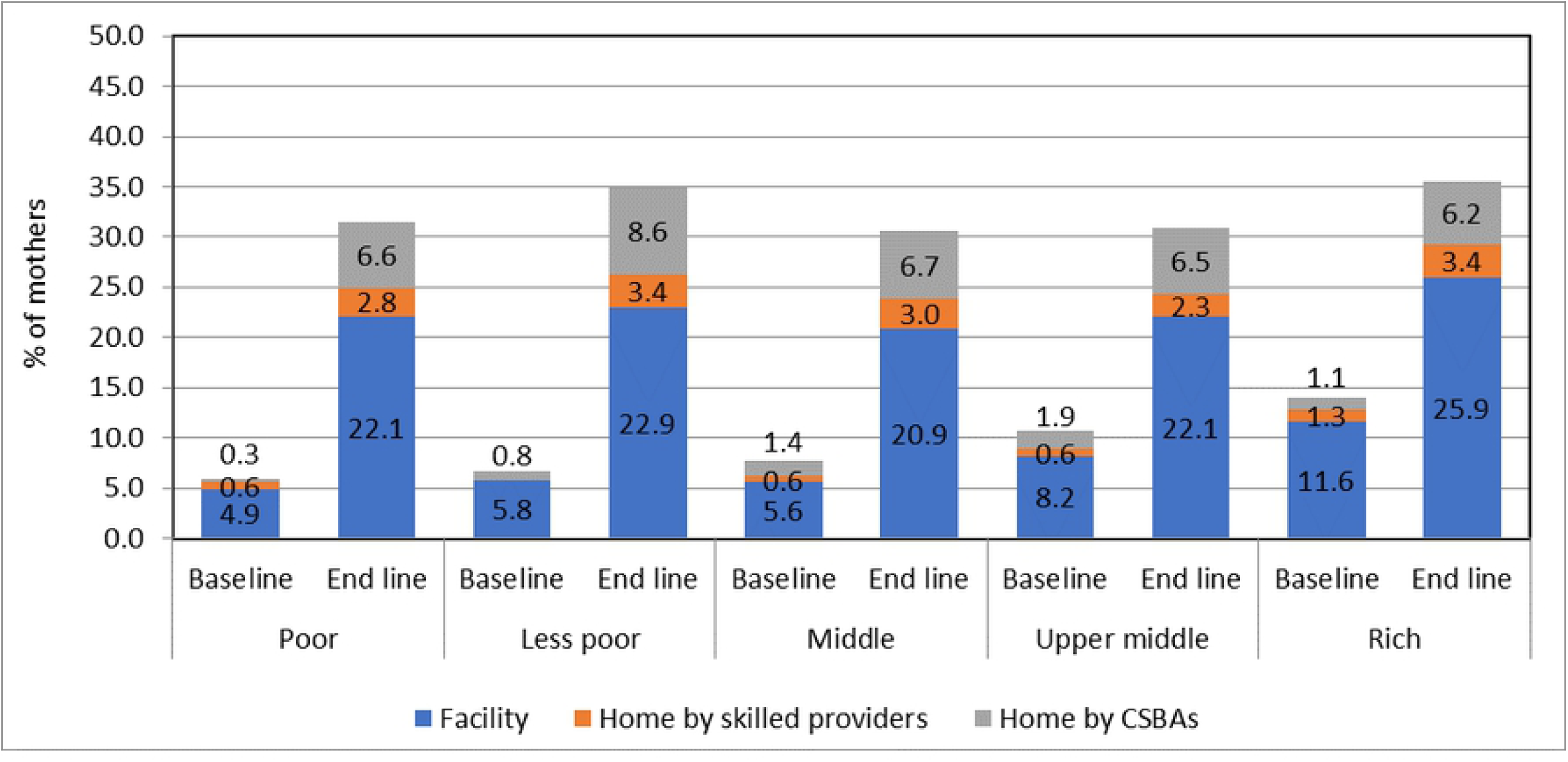
Source of skilled 4+ ANC by socioeconomic status during the baseline and endline survey.

**“Figure 2”** describes despite the other sources of birth attendants, mothers from all wealth groups inclined to have CSBAs as compared to either doctors or nurses/FWVs to conduct their deliveries at the endline period.

**Figure 2:**
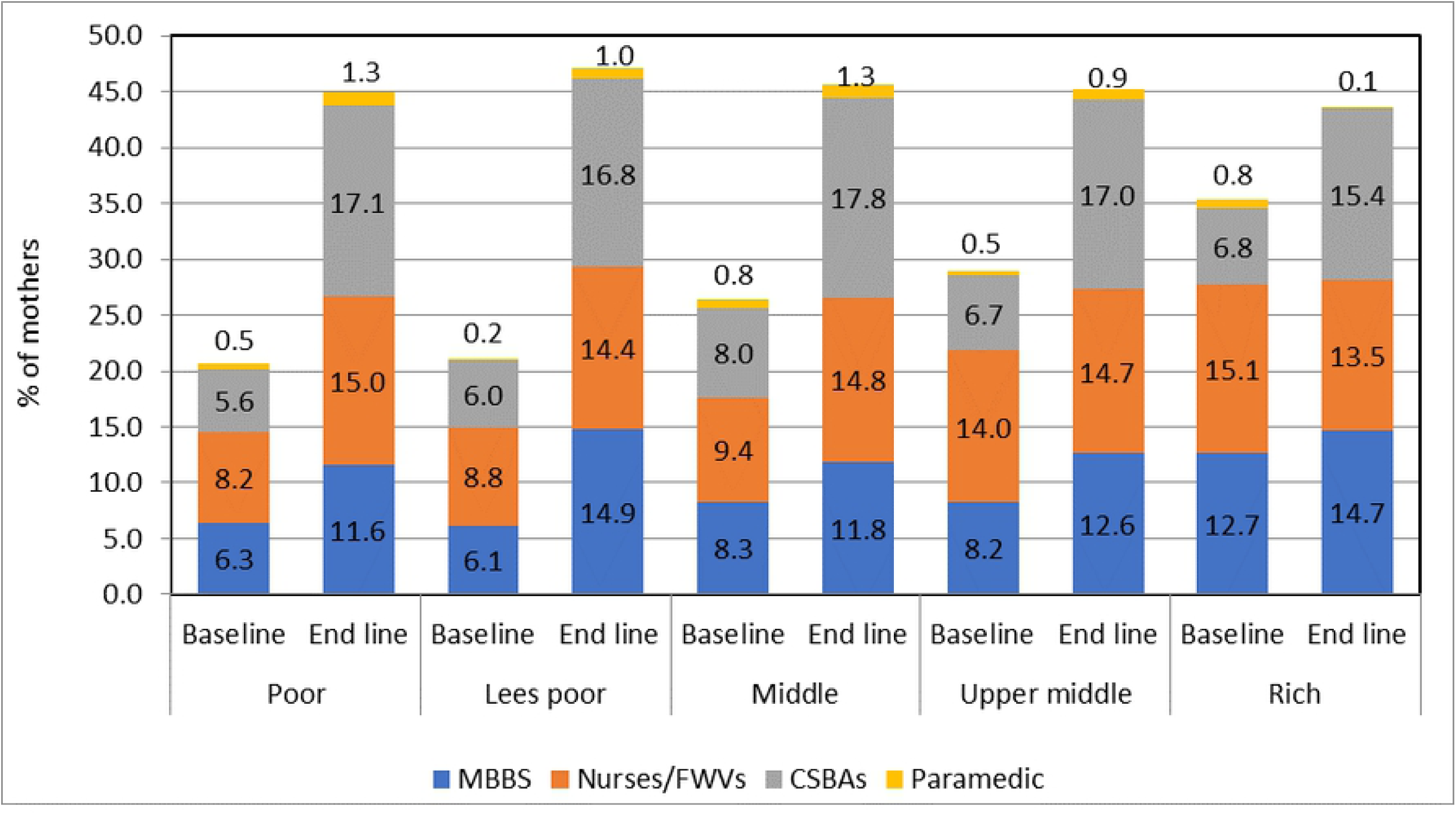
Source of skilled delivery by socioeconomic status during the baseline and endline survey.

**“Figure 3”** indicates the utilization of the facility was noteworthy in all the wealth index groups for PNC also. Receipt of the PNC from the CSBAs also increased at the endline period, interestingly with almost the same proportion across the groups of wealth index.

**Figure 3:**
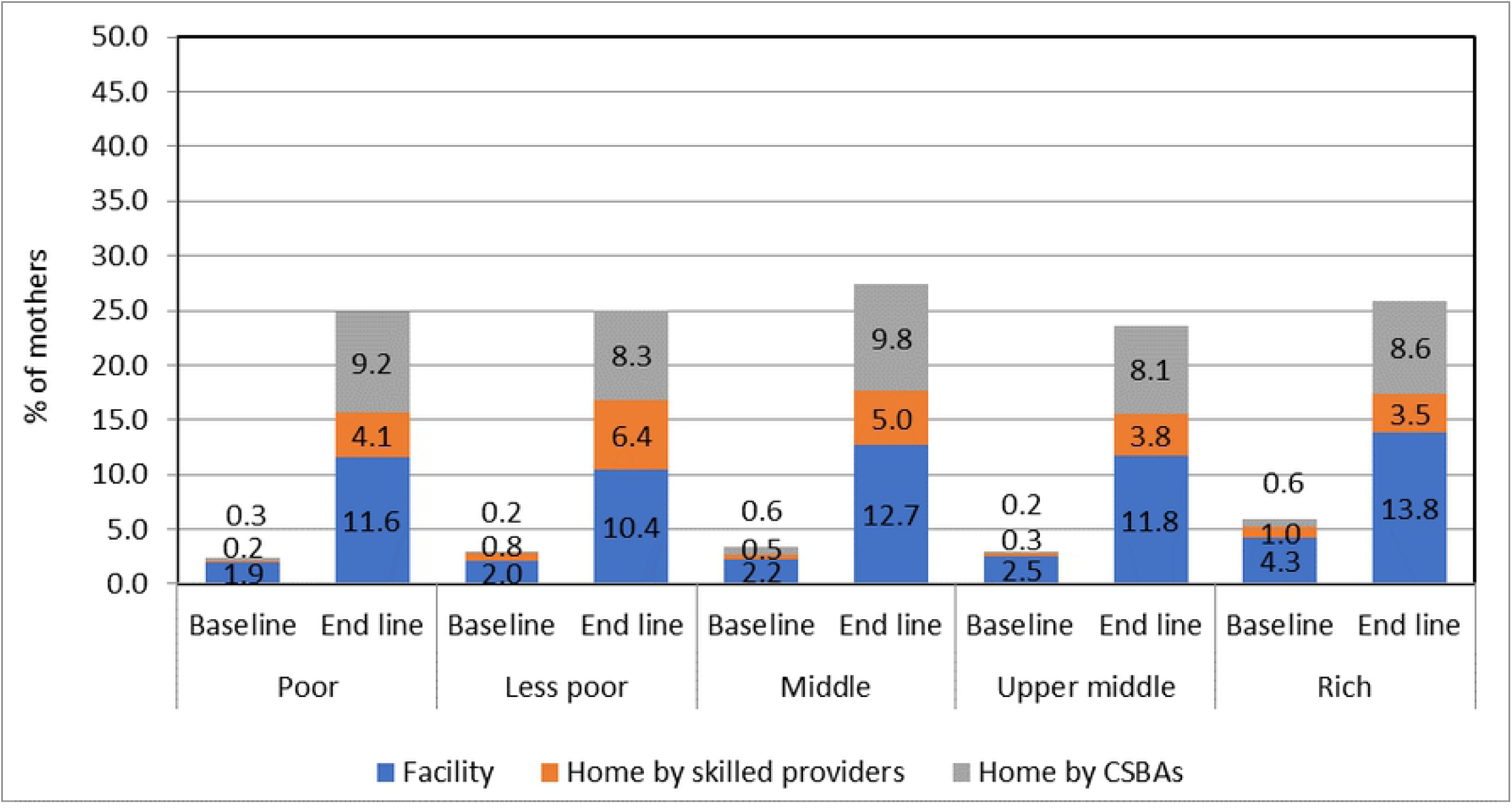
Source of skilled PNC by socioeconomic status during the baseline and endline survey.

**“Figure 4”** shows equity gap reduced between poor and rich in the endline for the services-ANC, childbirth and PNC in comparison to the baseline.

**Figure 4:**
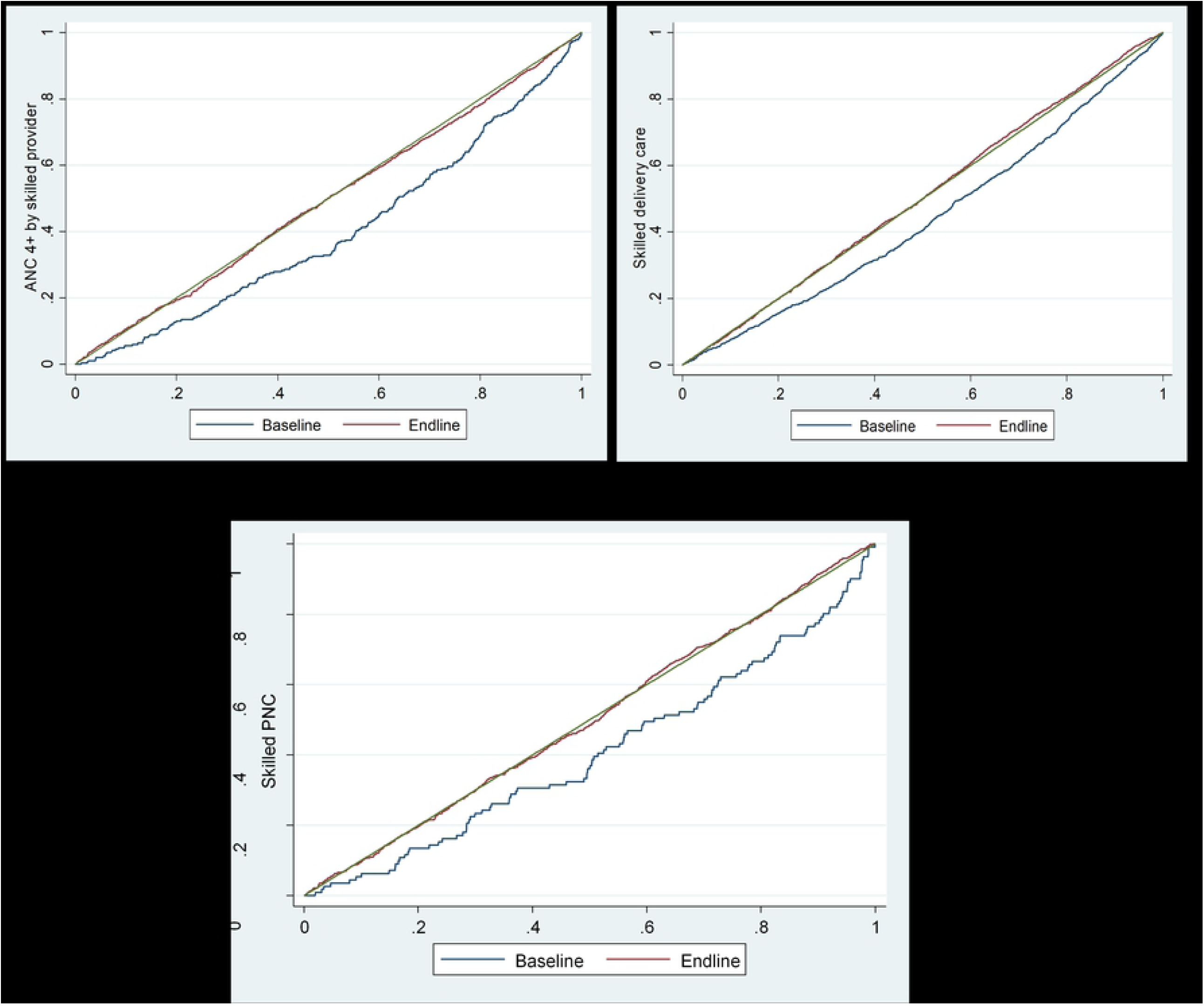
Concentration curve of skilled 4+ ANC, skilled delivery and skilled PNC utilization by study period.

**“Table 3”** shows the related concentration index decreased from 0.220 to 0.013 (p<0.001) for skilled 4+ANC utilization. The rich-poor ratio for skilled childbirth reduced from 1.7:1 to 1.0:1 and the related concentration index declined from 0.161 to -0.021 (p<0.001). A similar reduction was also observed in the utilization of skilled PNC; where the rich-poor gap decreased from 2.5:1 to 1:1 and the related concentration index declined from 0.197 to -0.004 (p<0.001). Figures 4, 5, and 6 showed the concentration curves for the utilization of skilled 4+ANC, childbirth, and PNC between the baseline and the endline periods.

**Table 3:** Measure of inequity (Concentration Index and 95% Confidence Interval) by study period.

| MNH care services | Baseline |  |  | Endline |  |  | % Change | p-value |
| --- | --- | --- | --- | --- | --- | --- | --- | --- |
|  | CI | LL | UL | CI | LL | UL |  |  |
| 4+ANC by skilled providers | 0.220 | 0.219 | 0.221 | 0.013 | 0.0125 | 0.0139 | -0.207 | <0.001 |
| Delivery by skilled providers | 0.161 | 0.159 | 0.161 | -0.021 | -0.021 | -0.020 | -0.181 | <0.001 |
| PNC by skilled providers | 0.197 | 0.177 | 0.215 | -0.004 | -0.012 | 0.003 | -0.201 | <0.001 |
| CI= Concentration Index<br>LL= Lower Limit<br>UL= Upper Limit |  |  |  |  |  |  |  |  |

## Discussion

This study finds that before providing any intervention, the inequity across the wealth index groups prevailed for all three pregnancy cares-ANC, delivery and PNC intensely. However, during endline, the difference resonated opportunities to achieve equity in improving skilled healthcare service by ensuring presence of skilled healthcare providers. Maternal health care during all the three pregnancy periods were better utilized by mothers of all five-wealth index group which contributes to the favorable results of the intervention. Other co-factors such as level of education, occupation and number of pregnancies played a very minimal role over the utilization of the skilled care across all the wealth groups as no significant difference is observed from the baseline to the endline.

Key factors constraining seeking facility care perhaps due to the inequities in the health facility coverage by geographic area, which did not entitle the poor. Different maternal health care services have been provided through district hospitals, upazila health center, upazila health and family welfare centers, and community clinics, since the 1980s by the Government of Bangladesh (19). But the service delivery module did not guide to reach the poor which recently has been changed through the attempt to ensuring universal health coverage for all (20). This study was proposed based on the inequity found in the MNCH care in 2007 which is prevailing till 2017. The series of Bangladesh Demographic and Health Survey measured that a big chunk of pregnant women of the poorest quintile doesn’t seek ANC from a medically trained provider and while almost all the pregnant women of the richest quintile receive ANC (7, 21-23). Further study suggests, half of the pregnant women in Bangladesh give birth at home and the majority is conducted by unskilled birth attendants. Furthermore, references discussed in background of this study talks for the long drawn inequity among the wealth groups on the health facility delivery and uptake of PNC (7). Hence, specific actions for the governments include initiation of actions to provide universal coverage of essential interventions and packages of service delivery prioritizing availability of skilled health care providers should be emphasized.

Connecting obstetric providers to the women of the poor groups impacted on the increased facility maternity care (24). This study put attention to the impact of the even distribution of CSBAs across the different wealth index groups and their respective physical remoteness, where scarcity of the skilled providers is an obstacle for the basic and comprehensive emergency obstetric care We found that the integrated intervention could make significant progress in skilled maternal health care utilization in the selected sub-district of Bangladesh. All the key indicators of the utilization of skilled maternal care improved in this area over the 4-years implementation of the intervention package. Skilled ANC was increased several folds, skilled delivery care doubled among the poor groups and similarly, a robust improvement in skilled PNC was observed among the poor women. The integrated intervention package focusing on coverage of skilled healthcare providers like CSBAs is assumed to provide opportunities for women to be informed on the necessity of the facility care as well. A similar intervention showed to have positive effects on the skilled provider’s care during pregnancy and delivery in remote areas by poor mothers (12).

The ANC is rationalized to maintain the continuum of care (25), for example, facilitation of mother’s access to skilled delivery and postpartum care (26). One of the important findings of this study was a decreased rich-poor gap for the skilled ANC services at the end of the intervention. The intervention was successful in improving equitable access to the ANC services that reflected in the smaller gaps between rich and poor for availing 4+ ANC from skilled providers. Provision of skilled ANC is expected to encourage women, especially the poor to seek skilled care during delivery and after delivery (PNC). This might prevent the practice of seeking care at the height of the maternal emergency, thus saving money for advanced management as well as a woman’s precious life. Ultimately, these implications would end-up with a cost-effective benefit of improved maternity services on mortality and morbidity reduction (25). Although, the argument presented in the association of increased ANC visits with maternal mortality reduction(27), maternity services including ANC by the local midwives and efficient referral systems showed a reduction in maternal mortality (28). Despite the reduction of the rich-poor gap and the significant improvement in 4+ANC by the poor, the 4+ANC rate from the skilled providers remained considerably low among them (around one-third) in the sub-district at the endline. Therefore, greater efforts are needed in obtaining better access to skilled ANC services by the poor. However, a significant increase in the utilization of the facility for ANC services at the endline would be conclusive about the effect of the intervention.

Although significant changes in using skilled delivery care had been marked between the baseline and the endline across the five wealth index categories, overall utilization of skilled birth attendants was doubled at the end of the intervention among the poor women that equalized the gap of the baseline to that of the rich quintile; apparently, CSBAs’ proportionate coverage by the intervention among all the quintiles might effect in reducing such gaps, respective of the quintiles’ significant increased utilization of CSBAs during delivery. Facility delivery was also doubled by the poor women after the intervention, this might be due to the associated awareness components of the intervention, such as counseling by CSBAs, CSGs, the orientation of the providers of the primary level health facilities. That stimulated an efficient follow-up of a fundamental second option of facility delivery for more effective intrapartum strategies (29). It is encouraged to conduct delivery at facility especially for the high-risk women; because use of a skilled attendant at home needs more complex logistic issues, for example, recognition of initial immediate management of complications and cost-effective transport mechanism for the compliance to their referral decision (30).

In Bangladesh, facility delivery remains low despite the improved healthcare delivery infrastructure (29, 30). Home-based intrapartum care preference should be advocated to replace with facility delivery to reduce the burden of recognition of complications and arrangement of transport on families. Facility maternity care placed many countries with maternal mortality less than 200 deaths or even lower per 100,000 live births (29). Nevertheless, facility-based care alone is not enough to reduce maternal mortality, rather, substantiating the significant decline in maternal mortality ratio (29). The MIRA study in Makwanpur, Nepal, concluded that simply strengthening health facilities is unlikely to influence perinatal care-seeking practices in a situation of preference to community-based management (26), as evidenced in another study that the poor women choose the nearby health facility even though the facility has compromised quality of care (31). Therefore, investment only in CSBAs is not justifiable for maternal mortality reduction without improving the utilization of the facility for delivery care and quality of the components.

Reduction in the risk of adverse maternal and neonatal outcomes is also associated with PNC timing (29). Skilled PNC has been increased to several folds at the endline; the significant decrease in the rich-poor gaps for skilled PNC services might be explained by an indirect effect of family value for improving newborn’s survival (32). However, this improvement remained to one-fourth in all wealth index groups at the endline, indicating a further emphasis on the PNC intervention strategies. Generally, PNC within 2 days following childbirth is very high for the facility delivery in comparison to the home delivery (7). Association between the facility delivery and the PNC was not measured but the descriptive statistics revealed that the facility-based PNC had been followed a similar trend like the facility delivery among the poor. As already mentioned this intervention did not take into full account the resources needed to achieve high uptake of skilled PNC that usually counted at the facility after childbirth, therefore, change in preference from home to facility delivery has also prospect to increase the optimal skilled PNC among the poor mothers.

## Strength and Limitation

This study is the first of its kind housing the most essential maternal health service components combined together as an integrated package to maximize the service quality related to maternal health in a remote setting as ours. Further, this study successfully employed one skilled provider per ten thousand population in community setting as per WHO health human resource guideline and ensured community engagement and the overall efforts translated into the improved maternal health service utilization in the endline than the baseline. However, this study left few rooms of improvements as this study evaluation did not measure cost, thus preventing the detection of its sustainability. This experimental research study observed the effect size based on a before-and-after design without comparing it with a control group. Therefore, it is difficult to exclude the possibility of others’ intervention effects.

## Conclusion

The significant improvements in skilled ANC, delivery care, and PNC by poor mothers in this study might have been influenced through implementation of the integrated demand- and supply-side interventions package at community level. On the contrary, since the significant increase in skilled maternity care remained low in this study that merely reminds the non-inclination to skilled providers by rural women. It is critical to have a proper blend of both the demand- and supply-side MNH interventions especially in one instance in Bangladesh. Further, research should focus on the community demands about the health system issues to overcome barriers to achieve progress. This study results indicate that the existing inequity in the health systems can be reduced by adopting the dual programmatic approaches involving both the community and the health facilities. In this intervention, establishing community support groups created community awareness that might support poor pregnant women to reach health facilities either for routine care or during emergencies. On the supply-side, this intervention equipped the skilled health workforce of the health facilities through refresher training on MNCH to ensure the quality of care to the mothers and their infants that help to build trust in the community.

## Data Availability

All relevant data are within the manuscript and its Supporting Information files.

## Acknowledgement

icddr, b acknowledges with gratitude the financial contribution of Australian Agency for International Development (AusAID) for funding this study. We are grateful to our study participants for their spontaneous participation and sincere commitment to fulfill the research endeavor. icddr, b is also grateful to the Government of Bangladesh, Canada, Sweden and the UK for providing core/ unrestricted support.

